# Shorter androgen receptor polyQ alleles protect against life-threatening COVID-19 disease in males

**DOI:** 10.1101/2020.11.04.20225680

**Authors:** Margherita Baldassarri, Nicola Picchiotti, Francesca Fava, Chiara Fallerini, Elisa Benetti, Sergio Daga, Floriana Valentino, Gabriella Doddato, Simone Furini, Annarita Giliberti, Rossella Tita, Sara Amitrano, Mirella Bruttini, Susanna Croci, Ilaria Meloni, Anna Maria Pinto, Chiara Gabbi, Francesca Sciarra, Mary Anna Venneri, Marco Gori, Maurizio Sanarico, Francis P. Crawley, Uberto Pagotto, Flaminia Fanelli, Marco Mezzullo, Elena Dominguez-Garrido, Laura Planas-Serra, Agatha Schlüter, Roger Colobran, Pere Soler-Palacin, Pablo Lapunzina, Jair Tenorio, Spanish Covid HGE, Aurora Pujol, Maria Grazia Castagna, Marco Marcelli, Andrea M. Isidori, GEN-COVID Multicenter Study, Alessandra Renieri, Elisa Frullanti, Francesca Mari

## Abstract

**Background:** COVID-19 presentation ranges from asymptomatic to fatal. The variability in severity may be due in part to impaired Interferon type I response due to specific mutations in the host genome or to autoantibodies, explaining about 15% of the cases when combined. Exploring the host genome is thus warranted to further elucidate disease variability.

**Methods:** We developed a synthetic approach to genetic data representation using machine learning methods to investigate complementary genetic variability in COVID-19 infected patients that may explain disease severity, due to poly-amino acids repeat polymorphisms. Using host whole-exome sequencing data, we compared extreme phenotypic presentations (338 severe versus 300 asymptomatic cases) of the entire (men and women) Italian GEN-COVID cohort of 1178 subjects infected with SARS-CoV-2. We then applied the LASSO Logistic Regression model on Boolean gene-based representation of the poly-amino acids variability.

**Findings:** Shorter polyQ alleles (≤22) in the androgen receptor (*AR*) conferred protection against a more severe outcome in COVID-19 infection. In the subgroup of males with age <60 years, testosterone was higher in subjects with *AR* long-polyQ (≥23), possibly indicating receptor resistance (p=0.004 Mann-Whitney U test). Inappropriately low testosterone levels for the long-polyQ alleles predicted the need for intensive care in COVID-19 infected men. In agreement with the known anti-inflammatory action of testosterone, patients with long-polyQ (≥23) and age>60 years had increased levels of C Reactive Protein (p=0.018).

**Interpretation:** Our results may contribute to design reliable clinical and public health measures and provide a rationale to test testosterone treatment as adjuvant therapy in symptomatic COVID-19 men expressing *AR* polyQ longer than 23 repeats.

**Funding:** MIUR project “Dipartimenti di Eccellenza 2018-2020” to Department of Medical Biotechnologies University of Siena, Italy (Italian D.L. n.18 March 17, 2020). Private donors for COVID research and charity funds from Intesa San Paolo.

**Boxes:** *Evidence before this study:* We searched on Medline, EMBASE, and Pubmed for articles published from January 2020 to August 2020 using various combinations of the search terms “sex-difference”, “gender” AND SARS-Cov-2, or COVID. Epidemiological studies indicate that men and women are similarly infected by COVID-19, but the outcome is less favorable in men, independently of age. Several studies also showed that patients with hypogonadism tend to be more severely affected. A prompt intervention directed toward the most fragile subjects with SARS-Cov2 infection is currently the only strategy to reduce mortality. glucocorticoid treatment has been found cost-effective in improving the outcome of severe cases. Clinical algorithms have been proposed, but little is known on the ability of genetic profiling to predict outcome and disclose novel therapeutic strategies.

*Added-value of this study:* In a cohort of 1178 men and women with COVID-19, we used a supervised machine learning approach on a synthetic representation of the uncovered variability of the human genome due to poly-amino acid repeats. Comparing the genotype of patients with extreme manifestations (severe vs. asymptomatic), we found that the poly-glutamine repeat of the androgen receptor (AR) gene is relevant for COVID-19 disease and defective AR signaling identifies an association between male sex, testosterone exposure, and COVID-19 outcome. Failure of the endocrine feedback to overcome AR signaling defect by increasing testosterone levels during the infection leads to the fact that polyQ becomes dominant to T levels for the clinical outcome.

*Implications of all the available evidence:* We identify the first genetic polymorphism predisposing some men to develop a more severe disease irrespectively of age. Based on this, we suggest that sizing the *AR* poly-glutamine repeat has important implications in the diagnostic pipeline of patients affected by life-threatening COVID-19 infection. Most importantly, our studies open to the potential of using testosterone as adjuvant therapy for severe COVID-19 patients having defective androgen signaling, defined by this study as ≥23 PolyQ repeats and inappropriate levels of circulating androgens.

## INTRODUCTION

This study was based on a preliminary descriptive analysis of a cohort of 35 hospitalized COVID-19 patients that was used to pinpoint a combined model of rare and common host genetic variants.^1^ After this initial study, we extended the cohort to more than 1,000 Italian SARS-CoV-2 PCR-positive subjects.^2^ Among them, 25% were asymptomatic, 25% were very severe, and the remaining presented with different levels of respiratory failure and multi-organ involvement.^2^

Alongside the mode of transmission, viral load, comorbidities, and demographic factors (such as age and sex), the host genetic background appears to play an important role in COVID-19 severity and progression.^3^ Data from classical methods of analysis, such as Genome-Wide Association Study (GWAS) or burden gene test on whole-exome sequencing (WES) data, have pinpointed a limited number of genetic factors that do not entirely explain how the host’s genetic background contributes to COVID-19 severity^4^. Two groups have recently identified rare variants in the interferon type I pathway that are responsible for inborn errors of immunity in a small percentage of patients and auto-antibodies against type I interferon genes in up to 10% of with severe COVID-19^5,6,7^. We hypothesized that most patients might instead have a more complex genetic background consisting of mutations arising in common susceptibility genes where additional private, rare, or low-frequency mutations provide the virus with the most favorable environment for replication, spread, and organ damage. Thus, we have considered the possibility that poly-amino acids repeat polymorphisms may contribute to COVID-19 severity. Our hypothesis was tested using a method of analysis based on a synthetic approach to genetic data representation. Specifically, we considered that polymorphisms in the polyQ tract of the Androgen Receptor (*AR*) may protect against the development of severe COVID-19. AR contains in its N-terminus domain a polymorphic polyQ tract, ranging between 9 and 36 repeated CAG units in the normal population^8^. *In vitro* and *in vivo* studies have demonstrated that the transactivation potential of AR is inversely correlated to repeat length and Q-tract length can significantly influence androgen-dependent physiological processes^8-11^.

Several lines of evidence lead to the concept that androgens are relevant to both SARS-CoV-2 infection and COVID-19 disease presentation; however, they seem to have a Janus bifacial way of action^12,13^. On one side, androgens promote the transcription of the *TMPRSS2* gene that encodes a serine protease known to prime the spike (S) protein of coronaviruses, facilitating viral entry into the cells^14^. On the other hand, hypogonadism and not hypergonadism is known to correlate with severe COVID-19^15^ and other chronic conditions, partly due to the loss of attenuation of the inflammatory immune response by testosterone^16-18^. Functional hypogonadism is also very common in male obesity and diabetes mellitus, two recognized factors of poorer prognosis in COVID-19 patients. The synthetic supervised genetic approach reported here contributes to disentangling this complex androgen-COVID-19 relationship. It may also have significant implications for further development of precision medicine in COVID-19 and identifies the use of androgens as a potential way to shape disease outcome as earlier proposed for glucocorticoids^19^, and subsequently demonstrated in a clinical trial^20^.

## RESULTS

### Testing the role of common poly-amino acid repeat polymorphisms in COVID-19 outcome

In order to test the role of common poly-amino acid repeat polymorphisms in determining COVID-19 clinical severity, we performed a nested case-control study (NCC), selecting the extreme phenotypic ends of our entire GENCOVID cohort (**Table 1**). (**Figure 1**). Among 18,439 annotated genes we selected those with amino acid repeats, namely 43 genes, and represented them as a boolean. Logistic regression with LASSO regularization analysis identified *AR* as the only protective gene (**Figure 1, panel A**). As expected, the grid search curve of the cross-validation score (**Figure 1, panel C**) shows a maximum for an intermediate value of the L1 regularization parameters, and the chosen parameter is 6.31. With this calibration setting, the 10-fold cross-validation provides good performances in terms of accuracy (77%), precision (81%), sensitivity (77%), and specificity (78%) as shown in **Figure 1, panel D**. The confusion matrix is reported in **Figure 1, panel B**, whereas the Receiver Operating Characteristic (ROC) curve (**Figure 1, panel E**) provides an Area Under the Curve (AUC) score of 86%.

**Table 1.** Demographics characteristics of the Italian GEN-COVID Cohort and NCC study.

|  |  | **Intubation** | **CPAP/BiPAP**<br>**Ventilation** | **Oxygen Therapy** | **No respiratory**<br>**support** | **Oligo-**<br>**asymptomatics**<br>**without**<br>**hospitalization** |
| --- | --- | --- | --- | --- | --- | --- |
| **GEN-COVID** | Number of Subjects | 108 | 230 | 352 | 188 | 300 |
|  | Male/Female | 80/28 | 157/73 | 208/144 | 104/84 | 116/184 |
|  | Age males (years) | 61,52±11,43 | 62,75±13,48 | 63,41±14,53 | 55,99±15,44 | 47,40±13,23 |
|  | Age females (years) | 63,71±13,96 | 66,23±15,25 | 68,40±14,74 | 52,88±16,39 | 48,61±11,06 |
|  |  | **Cases** |  |  |  | **Controls** |
| **NCC study** | Number of Subjects | 338 |  |  |  | 300 |
|  | Male/Female | 237/101 |  |  |  | 116/184 |
|  | Age males (years) | 62,34±12,84 |  |  |  | 47,40±13,23 |
|  | Age females (years) | 65,53±14,94 |  |  |  | 48,61±11,06 |

**Figure 1.**
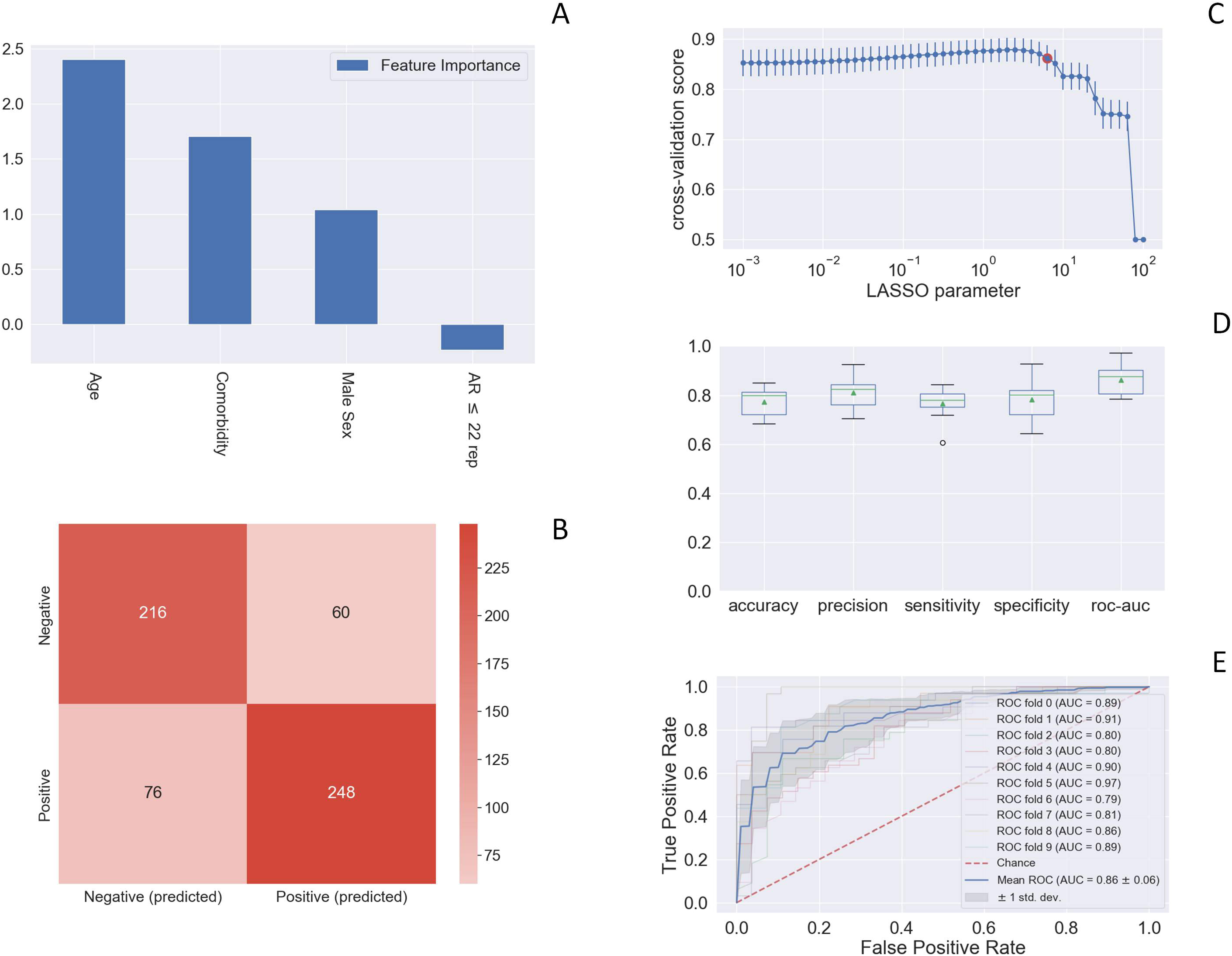
LASSO logistic regression. This histogram of the LASSO logistic regression weights represents the importance of each feature for the classification task (**Figure 1**) (**Panel A**). The positive weights reflect a susceptible behaviour of the features to the target COVID-19 disease, whereas the negative weights a protective action. The calculated odd ratio of AR short repeats (≤22) is 0,79 i.e. protective. Therefore, the odd ratio of long repeats (≥23) is 1/0,79= 1,27 i.e. severity. **Panel B**: Confusion matrix for the aggregation of the logistic regression predictions in the 10 folds of the cross-validation. **Panel C**: Cross-validation ROC-AUC score for the grid of LASSO regularization parameters; the error bar is given by the standard deviation of the score within the 10 folds; the red point corresponds to the parameter chosen for the fitting procedure. **Panel D**: Boxplot of accuracy, precision, sensitivity, specificity, and ROC-AUC score for the 10-fold of the cross-validation. The box extends from the Q1 to Q3 quartile, with a line at the median (Q2) and a triangle for the average. **Panel E**: ROC curve for the 10 folds of the cross-validation.

### Validation of polyQ polymorphism by sizing the PolyQ repeat of the *AR* receptor gene

In order to validate the results on *AR* obtained by LASSO logistic regression, we seized the male subset (351 subjects) using the gold standard technique that uses a fluorescent PCR reaction followed by the use of GeneScan Analysis software^®^ (Applied Biosystems) for sizing the number of triplets^21^. Based on the *AR* polyQ length, male patients were subdivided into two categories, those having a number of PolyQ repeats less than or equal to 22 repeats, and those having a number of PolyQ repeats greater than or equal to 23 repeats, being 23 repeats the reference sequence on genome browsers. We found that PolyQ repeats below 22 are enriched in the asymptomatic cohort of males. The difference was statistically significant in the group of males younger than 60 years of age in which genetic factors are expected to have a major impact (p-value 0,024) (**Table 2; Supplementary Table S1; Supplementary Table S2)**.

**Table 2.** PolyQ alleles correlation with COVID-19 outcome - males with age <60.

| **Males <60** |  |  |  |
| --- | --- | --- | --- |
|  | **<22** | **>23** | ***Marginal Row Totals*** |
| **Cases** | 52 (59,15%) | 36 (28,85%) | 88(48,08%) |
| **Controls** | 71 (63,85%) | 24 (31,15%) | 95(51,91%) |
| ***Marginal Column Totals*** | 123 (67,21%) | 60 (32,78%) | 183 (Grand Total) |

\* p-value (cases vs controls) = 0,024

### Validation of polyQ polymorphism in the Spanish Cohort

We then seized the polyQ repeat in an independent cohort of 158 males from Spain, with age < 60 years and without known comorbidities (117 cases and 41 controls). The association with shorter repeats (≤ 22) and protection was confirmed (p-value 0,014139 by χ^2^ test) (**Table 3**).

**Table 3.** Validation in Spanish cohort.

| Spanish validation ($\chi^2$) | | | |
| --- | --- | --- | --- |
| Males global |  |  |  |
| | $\leq 22$ | $\geq 23$ | *Marginal Row Totals* |
| **Cases** | 51 (32,27%) | 66 (41,77%) | 117 (74,05%) |
| **Controls** | 27 (17,08%)* | 14 (8,86%) | 41 (25,94%) |
| *Marginal Column Totals* | 78 (49,36%) | 80 (50,63%) | 158 (Grand Total) |

\* p-value (cases vs controls) = 0,014139 (Significant at $p < 0.05$ )

### Males with longer polyQ have receptor resistance

To functionally link the length of the PolyQ repeats to AR functionality, we measured total testosterone (TT) in 183 men using LCMS/MS (**Supplementary Table S1**). TT was higher in patients carrying ≥23 vs ≤22 glutamines (13.45 vs 11.23 nmol/L, p-value of 0.0422), reflecting reduced negative feedback from the less active receptors present in patients carrying a PolyQ repeat of ≥23 (**Fig. 2a**).

**Figure 2.**
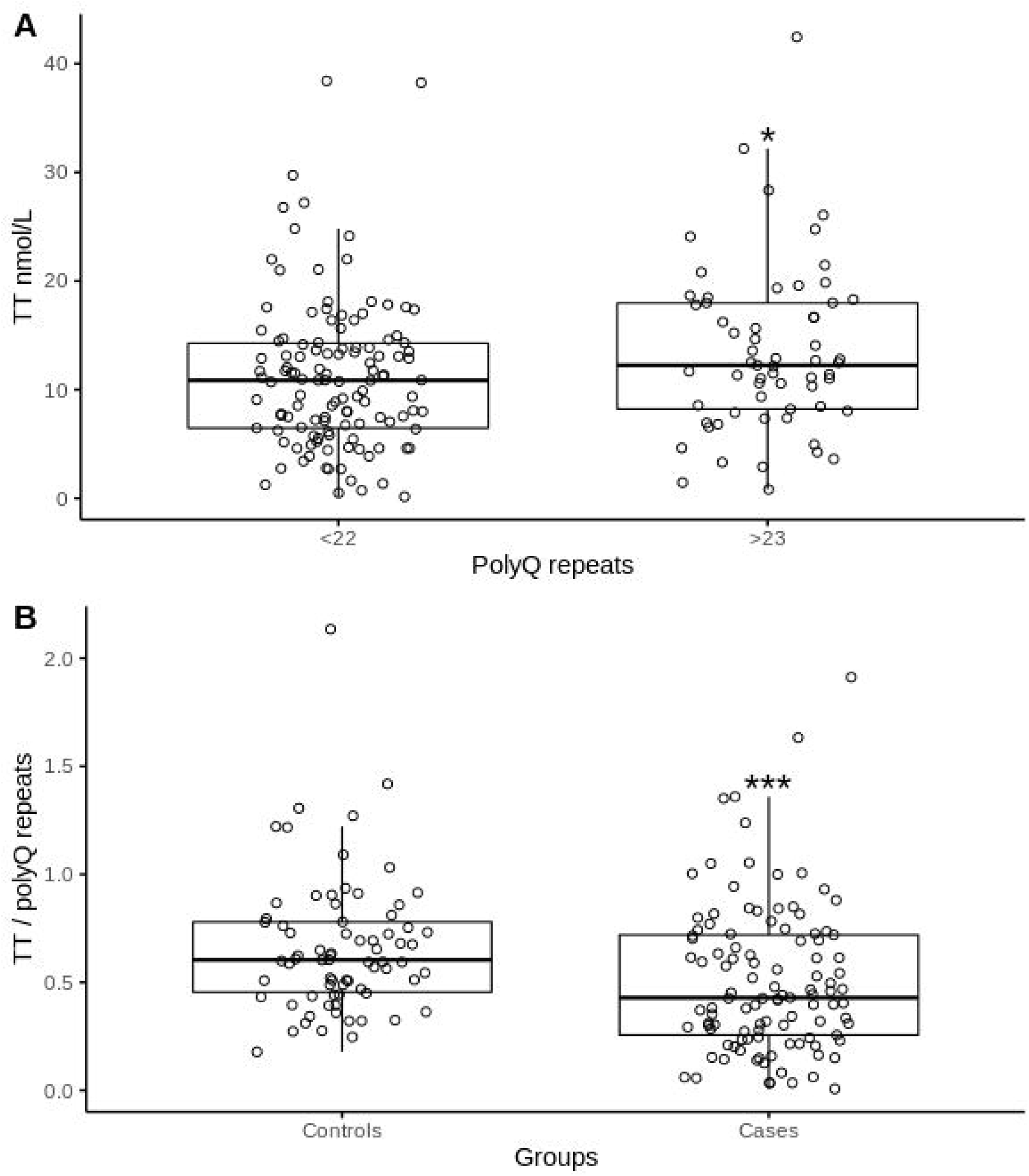
Relationship between Total Testosterone and polyQ repeats and clinical outcome. **Panel A:** Increased Total Testosterone levels in patients with longer polyQ repeats. Box-plot showing values of Total Testosterone (TT), expressed in nmol/L, in patients with shorter (≤22) and longer (≥23) polyQ repeats in AR gene. A subset of 183 males is studied: 122 males with polyQ repeats ≤22 and 61 with polyQ repeats ≥23. The TT median value, represented by the black horizontal line, is higher in patients with ≥23 polyQ repeats. (*p-value = 0.0422; Mann-Whitney U test). **Panel B:** Reduced Testosterone/PolyQ ratio in patients with the most severe COVID-19 clinical presentation. Box-plot showing the ratio Total Testosterone (TT), over the number of polyQ repeats, in cases and controls. A subset of 183 males (109 cases, 74 controls) was studied. The median of TT Ratio (nmol/L/polyQ number), represented by the black horizontal line, is lower in patients that underwent intubation/CPAP/BiPAP ventilation (cases). (***p-valu = 0,00038; Mann-Whitney U test).

### Unbalanced Testosterone-AR axis in males with longer polyQ repeats

Hormonal status of the entire male set sized for polyQ according to the age group revealed lower TT and calculated free T levels and higher SHBG levels with increasing age (**Supplementary Table S3**).

To evaluate whether the AR receptor reduced activity resulted in signs and symptoms of hypogonadism, subjects were interviewed, post-infection, using a modified version of the Androstest®^22^. Interviews were available for 61 subjects (representative of the extremes genotypes of the cohort: 43 with ≤19 repeats and 18 with ≥25 repeats). An Androtest score ≥8 was found in 38% of men with ≥25 repeats as compared to 16% of those with ≤19 glutamines (likelihood ratio, p=0.046). Similarly, cryptorchidism (11% vs. 2%), anemia (11% vs 2%) and severe erectile dysfunction (22% vs. 9%) were more frequently reported in subjects with ≥25 repeats, but not osteopenia/osteoporosis (6% vs 7%). These results indicate a trend toward clinical hypogonadism for those with longer repeats. Conversely, in the entire male dataset, 6 cases of prostate cancer were found annotated in the past-medical history, all in the ≤22 glutamines group, suggesting an increased prostate sensitivity to androgens. No difference was found in the prevalence of BPH or 5-alfa-reductase inhibitors use.

As the reduced signal transduction of AR might be compensated by higher testosterone levels, as shown in **Fig 2a**, we tested whether the feedback was sufficient to overcome polyQ repeats. A binomial logistic regression was performed to ascertain the interaction between age, testosterone levels and polyglutamine receptor length on the likelihood that subjects require intensive care during COVID infections. The logistic regression model was highly significant (χ^2^ (3) = 18,881, p<0.0001), with the model explaining 7.5% (Nagelkerke /R2) of the variance in COVID19 outcome **(Supplementary Table S4)**. A significant testosterone by polyglutamine length interaction was found (p=0.018), suggesting impaired feedback as predictor of the worst outcome, namely intubation or CPAP/BiPAP versus hospitalization not requiring respiratory assistance. To provide a graphical representation, we plotted the ratio between TT serum concentrations and polyQ number vs. clinical outcome (**Fig. 2b**). Results show a decreased mean ratio, a sign of an inappropriate rise of TT for increasing polyglutamine repeats, was associated with a worse outcome (p=0.00038).

### Older severely affected males with longer polyQ repeats display a more pronounced inflammatory phenotype

Finally, we tested the relationship between the AR polyQ repeat size and 5 laboratory markers of immunity/inflammation, including CRP, Fibrinogen, IL6, CD4 and NK count. We found that older (≥60) males with AR polyQ tract ≥23 have a higher (55.92 versus 48.21 mg/dl) mean value of CRP (p=0.018) and lower mean value of Fibrinogen and a trend of higher IL6 (**Table 4)**.

**Table 4.** Correlation between polyQ repeats in AR gene and laboratory values.

**CRP M≥60y cases**
| Triplets | Mean | Count | p-value |
| --- | --- | --- | --- |
| ≤22 | 48,21 | 78 | 0,01804 (Significant at p<0,05) |
| ≥23 | 55,92 | 38 |  |

**Fibrinogen M≥60y cases**
| Triplets | Mean | Count | p-value |
| --- | --- | --- | --- |
| ≤22 | 401,33 | 57 | 0,09277 |
| ≥23 | 320,34 | 27 |  |

**IL6 times the upper limit of normal M≥60y cases**
| Triplets | Mean | Count | p-value |
| --- | --- | --- | --- |
| ≤22 | 54,56 | 40 | 0,249 |
| ≥23 | 75,78 | 16 |  |

**CD4 Lymphocytes M≥60y cases**
| Triplets | Mean | Count | p-value |
| --- | --- | --- | --- |
| ≤22 | 264,06 | 32 | 0,22 |
| ≥23 | 357,52 | 21 |  |

**NK Cells M≥60y cases**
| Triplets | Mean | Count | p-value |
| --- | --- | --- | --- |
| ≤22 | 70,71 | 28 | 0,179 |
| ≥23 | 102,25 | 16 |  |

**CRP M<60y cases**
| Triplets | Mean | Count | p-value |
| --- | --- | --- | --- |
| ≤22 | 54,5 | 43 | 0,2 |
| ≥23 | 26,41 | 29 |  |

**Fibrinogen M<60y cases**
| Triplets | Mean | Count | p-value |
| --- | --- | --- | --- |
| ≤22 | 316,93 | 22 | 0,53 |
| ≥23 | 356,91 | 19 |  |

**IL6 times the upper limit of normal M<60y cases**
| Triplets | Mean | Count | p-value |
| --- | --- | --- | --- |
| ≤22 | 40,43 | 17 | 0,81 |
| ≥23 | 31,8 | 14 |  |

**CD4 Lymphocytes M<60 cases**
| Triplets | Mean | Count | p-value |
| --- | --- | --- | --- |
| ≤22 | 503,68 | 16 | 0,45 |
| ≥23 | 396,13 | 15 |  |

**NK Cells M<60y cases**
| Triplets | Mean | Count | p-value |
| --- | --- | --- | --- |
| ≤22 | 147,3 | 13 | 0,098 |
| ≥23 | 107,14 | 14 |  |

## DISCUSSION

We employed machine learning methodologies to identify a set of genes involved in the severity of COVID-19. In the presence of very high dimensionality, as for instance in a WES study, it is crucial to select the most predictive genes representing patterns of variation (mutations or variants) in subjects with different classes of response (i.e., disease state: from asymptomatic to severe cases). This problem is even more complex in diseases where multiple genes are involved in determining the severity and clinical variability of the pathology. Here, we wanted to represent poly-amino acids repeat polymorphisms that are typically missed in classical GWAS analysis, which concentrates on bi-allelic polymorphisms.

We used a machine learning approach and logistic regression with a LASSO regularization to test if using such a simplified representation could lead to a reliable prediction of extreme clinical outcomes (asymptomatic versus severely affected). This approach enabled us to predict such clinical outcomes with 77% sensitivity.

*AR* contains a highly variable polyglutamine repeat (poly-Q) located in the N-terminal domain of the protein, spanning from 9 to 36 glutamine residues in the normal population^5^. AR polyQ length correlates with receptor functionality, with shorter polymorphic glutamine repeats typically associated with higher and longer PolyQ tracts with lower receptor activity.^5^ AR is expressed in both males and females, but the bioavailability of its ligands testosterone (T) and dihydrotestosterone (DHT) differs significantly, being much higher in males. As previous studies linked male hypogonadism to a poorer outcome we decided to focus on male patients and demonstrated that shorter polymorphic glutamine repeats (≤22) confer protection against life-threatening COVID-19 in a subpopulation of individuals with age <60 years.

We also confirmed the functional association between polyQ size and receptor activity. Specifically, we showed that longer polyQ size (≥23) is associated with higher serum T levels, suggestive of impaired negative feedback (p=0.0046 at Mann-Whitney U test) at the level of the hypothalamus and pituitary gland, therefore confirming the association between functional hypogonadism and severe COVID-19 infection.

As T is known to have an immunomodulatory activity attenuating inflammatory immune responses,^23-27^ we hypothesized that a long PolyQ repeat would lead to a pro-inflammatory status heralded by increased proinflammatory markers^18,28^ by conferring decreased AR transcriptional activity. Conversely, men with a more active receptor (short PolyQ tract) would be protected because they can tame the inflammatory response and increase survival regardless of serum T levels. We found that-CRP-, one of the main inflammatory markers, was higher in subjects with a long AR PolyQ tract. This observation not only is in line with the known anti-inflammatory function of testosterone, but also reinforces the functional importance of the AR PolyQ tract and its association with COVID-19 clinical outcome. Furthermore, this observation suggests that CRP is hierarchically more indicative than serum T level, which can be inappropriately normal and mask a functional hypogonadism in men with a long PolyQ repeat.

The allele distribution of the PolyQ repeat length varies among different populations, with the shortest in Africans, medium in Caucasians, and longest in Asians^29^. Interestingly, WHO data on mortality rates during the first pandemic wave indicated a higher fatality rate in China and Italy (https://covid19.who.int/)^30^ with respect to African. Hence, AR polyQ length variability could represent an explanation for the observed differences in death rate. Moreover, Africans seem to be more prone to infection^31^. This observation could be due to a more active AR receptor, leading to a higher expression of *TMPRSS2*, a protease essential for SARS-CoV-2 spread^14^.

Different studies have shown an association between hypogonadism and severe COVID-19^15^ and other chronic obstructive pulmonary diseases^16,17^. Our results are in line with these initial observations and contribute to explain a possible mechanism leading to these associations. The present study brings the observations to the upper level, revealing that is the overall androgenic effect-resulting from the interaction of polyQ polymorphism and circulating testosterone levels- to predict the need for intensive care. In infected men, we observed impaired feedback no longer sufficient to compensate for the reduced AR transcriptional activity, leading to the fact that polyQ becomes dominant to T levels. The latter helps to solve some inconsistencies, including the early reports of a slightly better outcome in prostate cancer patients-who tend to have lower polyQ, as in our cohort - when compared to other cancers.

An improvement in peak oxygen saturation in men receiving testosterone replacement therapy has been demonstrated in a randomized controlled trial^32^ and could be one of the mechanisms responsible for the observed protective effect of AR’s with shorter polyQ tract in COVID-19 patients. Thus, our study has important implications and suggests that androgen therapy could be tested as an adjuvant treatment in COVID-19-infected men with functional hypogonadism. Based on the evidence discussed in this paper, a proper randomized controlled trial (RCT) is warranted in COVID-19 male patients with signs of hypoandrogenism and longer AR polyQ tract to test the safety and efficacy of AR agonists, like testosterone, in improving outcome. A simple genetic test measuring the *AR* receptor PolyQ repeat in male patients could then be introduced to screen individuals more likely to benefit from testosterone therapy.

Variants of another X-linked gene, TLR7, have been associated with severe COVID-19 outcomes in young men^6^. In the 2 reported families, the rare TLR7 mutations segregated as a highly penetrant monogenic X-linked recessive trait. While variants in TLR7 gene are expected to account for a small number of severely affected cases, our findings involve a much larger number of subjects, as although the OR is relatively small, long polyQ alleles are relatively common (27%)^33^. Overall, X-linked genetic variants keep coming up as important for defining severe COVID-19 cases in males.

In conclusion, we present a method that can predict if subjects infected by SARS-CoV-2 are at risk for life-threatening complications. This approach has 77% accuracy, 81% precision, 77% sensitivity, and 78% specificity. Furthermore, we present evidence suggesting that a more active AR has the potential to confer protection against COVID-19 severity. If confirmed, these observations should be followed by properly conducted clinical trials studying if testosterone helps decrease morbidity and mortality in patients affected by the most severe forms of the disease. Finally, as shown by regression analysis, ORs ranges between 1.26 and 1.45, therefore the risk of carrying a longer AR is much smaller than other already known strong predictors such as age and sex, but still is highly significant, relatively common, and among the very few known genetic predictors of COVID19 outcome.

## METHODS

### Patients

We performed a nested case-control study (NCC). Cases and controls were drawn from the Italian GENCOVID ^2^ cohort of 1178 subjects infected with SARS-CoV-2 diagnosed either by RT-PCR on nasopharyngeal swab, or by serology test. ^2^ Demographic characteristics of patients enrolled in the cohort are summarized in Table 1 according to their clinical status. In particular, the cohort was subdivided into 5 groups, based on the type of the respiratory and medical support that was given to the patients: *i*. endotracheal intubation; *ii*, CPAP-biPAP ventilation; *iii*. oxygen administration; *iv*. hospitalization without respiratory support; *v*. no hospitalization either because asymptomatic or with minor symptoms of COVID-19 (mild fever, cough, sore throat, etc.). Subjects of the cohort were recruited by Italian hospitals and primary care clinics participating in the GENCOVID Multicenter study previously described ^2^.

In the current NCC study, cases were selected according to the following inclusion criteria: *i*. CPAP/biPAP ventilation (230 subjects); *ii*. endotracheal intubation (108 subjects). As controls, 300 subjects were selected using the sole criterion of not requiring hospitalization. Cases and controls show the extreme phenotypic presentations of the GENCOVID cohort. Exclusion criteria for both cases and controls were *i*. SARS-CoV-2 infection not confirmed by PCR; *ii*. not-caucasian ethnicity. Demographic characteristics of the subjects in the NCC study are summarized in Table 1.

### Analysis of triplets size in the AR locus

To establish allele sizes of the polymorphic triplet in the AR locus, we used the HUMARA assay with minor modifications^21^. Specifically, we performed a fluorescent PCR followed by capillary electrophoresis on an ABI3130 sequencer. Allele size was established using the Genescan Analysis software.

### Binary representation of WES data

In this paper, one of several synthetic approaches to genetic data variability representation is used: poly-amino acids repeat polymorphisms (Benetti et al., Topological Data Analysis on boolean representation of genome variability as a method for discovering the genetic bases of complex disorders. Paper in preparation). Reads were mapped to the hg19 reference genome by the Burrow-Wheeler aligner BWA. Variants calling was performed according to the GATK4 best practice guidelines. Namely, duplicates were first removed by MarkDuplicates, and base qualities were recalibrated using BaseRecalibration and ApplyBQSR. HaplotypeCaller was used to calculate Genomic VCF files for each sample, which were then used for multi-sample calling by GenomicDBImport and GenotypeGVCF. In order to improve the specificity-sensitivity balance, variants quality scores were calculated by VariantRecalibrator and ApplyVQSR, and only variants with estimated truth sensitivity above 99.9% were retained. Variants were annotated by ANNOVAR. WES data were represented in a binary mode on a gene-by-gene basis.

#### Representation of poly-amino acids triple repeats

A set of binary variables was included to correctly describe non-frameshift variants in gene portions with repeated triplets. A total of 43 genes with triplet repeat variability were taken from UniProtKB (https://www.uniprot.org/uniprot/?query=reviewed:yes%20keyword:%22Triplet%20repeat%20expansion%20%5BKW-0818%5D%22). For any of these genes a variant Dij was defined as equal to 1 if the i-th gene presented a deletion in the region characterized by repeated triplets. Non-informative genes, with either all 0 or all 1, are removed from the dataset.

### LASSO logistic regression

The problem of extracting knowledge on the most relevant genes involved in the classification tasks of COVID-19 disease can be interpreted in the classical framework of feature selection analysis. Due to the specificities of the problem (i.e., the complex classification task), we adopted the embedded method of the LASSO logistic regression model. By denoting with □_*k*_ the coefficients of the logistic regression and by lambda (λ) the strength of the regularization, the LASSO (Least Absolute Shrinkage and Selection Operator) regularization^34^ term of the loss, 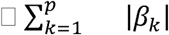, has the effect of shrinking the estimated coefficients to 0. This provides a feature selection method for sparse solutions within the classification tasks able to enforce both the sparsity and the interpretability of the results. The weights of the logistic regression algorithm can be interpreted as the feature importances of the subset of the most relevant features for the task^35^. The input features are the poly-amino acids triplet repeats presented in the previous section as well as gender, comorbidity (1 if there is at least one comorbidity) and the age, the latter as a continuous variable normalized between 0 and 1.

The fundamental hyper-parameter of the logistic regression algorithm is the strength of the LASSO term, which is tuned with a grid search method on the average area under the Receiver Operating Characteristic (ROC) curve for the 10-fold cross-validation. The regularization hyperparameter varies in the range [10^−3^, 10^2^] with 50 equally spaced values in the logarithmic scale. The optimal regularization parameter is chosen by selecting the most parsimonious parameter whose cross-validation score falls in the range of the best one along with its standard deviation. This choice is related to the aim of the work that is to select the most important genes (and not necessarily the entire set of genes contributing to COVID-19 variability). During the fitting procedure, the class slight unbalancing is tackled by penalizing the misclassification of the minority class with a multiplicative factor inversely proportional to the class frequencies. The data pre-processing was coded in Python, whereas for the logistic regression model the scikit-learn module with the liblinear coordinate descent optimization algorithm was used. Performances of the model were evaluated using the cross-validation confusion matrix as well as by computing precision, sensitivity, specificity, and the Receiver Operating Characteristic (ROC) curve.

### Total Testosterone measurement

Blood samples were collected after an overnight fast, immediately centrifuged at 4°C and stored at −20 °C until assayed. Blood samples were collected after an overnight fast, immediately centrifuged at 4 °C and stored at −20 °C until assayed. Serum total testosterone was measured using the Access testosterone assay (Beckman Coulter Inc., Fullerton, CA, USA) with a minimum detection limit of 0.35 nmol/L. Reference range for this assay was 6.07-27.1 nmol/L. Thawed plasma underwent 15 min incubation at 56°C for virus inactivation. Afterward, plasma total testosterone was measured by liquid chromatography - tandem mass spectrometry (LC-MS/MS) according to a previously validated method provided with reference values between 9.8 - 28.4 nmol/L^36^. In the present study, measurement was performed in 100 µl plasma, with sensitivity limit being 0.270 nmol/L, imprecision ranging 9.8 to 0.7% and accuracy 90.6 to 101.5% at concentration levels between 1.12 and 39.2 nmol/L. A stability test under viral inactivation conditions was performed in 6 samples, revealing a testosterone mean (min-max) % loss of 9.7% (4.6-16.7%). SHBG levels were measured in plasma samples using Quantikine ELISA Kit (DSHBG0B, R&D Systems, Minneapolis, MN, USA) according to the manufacturers’ instructions. Serum LH was measured using “Access LH assay “a chemiluminescent, two-step enzyme immunoassay (Beckman Coulter Inc., Fullerton, CA, USA). Sensitivity for the LH determination is 0.2 mIU/mL. Reference range in adult males for this assay is 1.2-8.6 mIU/mL

### Statistical analysis

Since serum and plasma Testosterone values were not normally distributed, the statistical analyses were performed using non-parametric tests. In particular, we used the Mann-Whitney U test to compare testosterone levels in males with AR long-polyQ (≥23) versus males with short polyQ (≤22). Logistic regression analysis was performed to ascertain the interaction between age, testosterone, and the number of polyglutamine repetitions and COVID19 outcomes. Box-Tidwell procedure was used to assess linearity and the Hosmer and Lemeshow to assess goodness of fit test.

## Data Availability

The data and samples referenced here are housed in the GEN-COVID Patient Registry and the GEN-COVID Biobank and are available for consultation. For further information, you may contact the corresponding author, Prof. Alessandra Renieri.

http://www.nig.cineca.it

https://www.covid19hg.org

## ACKNOWLEDGEMENTS

This study is part of the GEN-COVID Multicenter Study, https://sites.google.com/dbm.unisi.it/gen-covid, the Italian multicenter study aimed at identifying the COVID-19 host genetic bases. Specimens were provided by the COVID-19 Biobank of Siena, which is part of the Genetic Biobank of Siena, member of BBMRI-IT, of Telethon Network of Genetic Biobanks (project no. GTB18001), of EuroBioBank, and of RD-Connect. We thank the CINECA consortium for providing computational resources and the Network for Italian Genomes (NIG) http://www.nig.cineca.it for its support. We thank private donors for the support provided to A.R. (Department of Medical Biotechnologies, University of Siena) for the COVID-19 host genetics research project (D.L n.18 of March 17, 2020). We also thank the COVID-19 Host Genetics Initiative (https://www.covid19hg.org/), MIUR project “Dipartimenti di Eccellenza 2018-2020” to the Department of Medical Biotechnologies University of Siena, Italy. We also thank Intesa San Paolo for the 2020 charity fund dedicated to the project N. B/2020/0119 “Identificazione delle basi genetiche determinanti la variabilità clinica della risposta a COVID-19 nella popolazione italiana”.

## ETHICS APPROVAL

The GEN-COVID study was approved by the University Hospital of Siena Ethics Review Board (Protocol n. 16917, dated March 16, 2020). This observational study has been inserted in www.clinicaltrial.org (NCT04549831). The Spanish CovidHGE cohort is under IRB approval PR127/20 from Bellvitge University Hospital, Barcelona Spain.

## AUTHOR CONTRIBUTIONS STATEMENT

EF, FM, AR designed the study. CF and IM, were in charge of biological samples’ collection and biobanking. MB, FF were in charge of clinical data collection. MB, FF, AR, and FM performed analysis/interpretation of clinical data. UP, FF and MM performed testosterone measurement by LC-MS/MS. EDG, AS, AP and LPS performed the validation of association between shorter repeats and protection in a Spanish cohort. MM and AI critically reviewed the manuscript and interpreted clinical data/androgen physiopathological processes. SA and MB were in charge of DNA isolations from peripheral blood samples. FV, GD, AG, RT carried the sequencing experiments. EB, NP, SF, CG, MG and MS, performed bioinformatics and statistical analyses. EB, NP, SD, CF, and SC prepared Figures and Tables. EB, NP, AMP, FPC, AR, EF and FM wrote the manuscript. All authors have reviewed and approved the manuscript.

## ADDITIONAL INFORMATION

### GEN-COVID Multicenter Study (https://sites.google.com/dbm.unisi.it/gen-covid)

Francesca Montagnani^2,22^, Laura Di Sarno^1,2^, Andrea Tommasi^1,2,5^, Maria Palmieri^1,2^, Arianna Emiliozzi^2,22^, Massimiliano Fabbiani^22^, Barbara Rossetti^22^, Giacomo Zanelli^2,22^, Fausta Sestini^20^, Laura Bergantini^23^, Miriana D’Alessandro^23^, Paolo Cameli^23^, David Bennet^23^, Federico Anedda^24^, Simona Marcantonio^24^, Sabino Scolletta^24^, Federico Franchi^24^, Maria Antonietta Mazzei^25^, Susanna Guerrini^25^, Edoardo Conticini^26^, Luca Cantarini^26^, Bruno Frediani^26^, Danilo Tacconi^27^, Chiara Spertilli^27^, Marco Feri^28^, Alice Donati^28^, Raffaele Scala^29^, Luca Guidelli^29^, Genni Spargi^30^, Marta Corridi^30^, Cesira Nencioni^31^, Leonardo Croci^31^, Gian Piero Caldarelli^32^, Maurizio Spagnesi^33^, Paolo Piacentini^33^, Maria Bandini^33^, Elena Desanctis^33^, Silvia Cappelli^33^, Anna Canaccini^34^, Agnese Verzuri^34^, Valentina Anemoli^34^, Agostino Ognibene^35^, Massimo Vaghi^36^, Antonella D’Arminio Monforte^37^, Esther Merlini^37^, Federica Gaia Miraglia^37^, Mario U. Mondelli^38,39^, Stefania Mantovani^38^, Serena Ludovisi^38,39^, Massimo Girardis^40^, Sophie Venturelli^40^, Marco Sita^40^, Andrea Cossarizza^41^, Andrea Antinori^42^, Alessandra Vergori^42^, Stefano Rusconi^43,44^, Matteo Siano^44^, Arianna Gabrieli^44^, Agostino Riva^43,44^, Daniela Francisci^45,46^, Elisabetta Schiaroli^45^, Pier Giorgio Scotton^47^, Francesca Andretta^47^, Sandro Panese^48^, Stefano Baratti^48^ Renzo Scaggiante^49^, Francesca Gatti^49^, Saverio Giuseppe Parisi^50^, Francesco Castelli^51^, Eugenia Quiros-Roldan^51^, Melania Degli Antoni^51^, Isabella Zanella^52^, Matteo Della Monica^53^, Carmelo Piscopo^53^, Mario Capasso^54,55,56^, Roberta Russo^54,55^, Immacolata Andolfo^54,55^, Achille Iolascon^54,55^, Giuseppe Fiorentino^57^, Massimo Carella^58^, Marco Castori^58^, Giuseppe Merla^58^, Filippo Aucella^59^, Pamela Raggi^60^, Carmen Marciano^60^, Rita Perna^60^, Matteo Bassetti^61,62^, Antonio Di Biagio^62^, Maurizio Sanguinetti^63,64^, Luca Masucci^63,64^, Serafina Valente^65^, Maria Antonietta Mencarelli^5^, Caterina Lo Rizzo^5^, Elena Bargagli^23^, Marco Mandalà^66^, Alessia Giorli^66^, Lorenzo Salerni^66^, Patrizia Zucchi^67^, Pierpaolo Parravicini^67^, Elisabetta Menatti^68^, Tullio Trotta^69^, Ferdinando Giannattasio^69^, Gabriella Coiro^69^, Fabio Lena^70^, Domenico A. Coviello^71^, Cristina Mussini^72^, Giancarlo Bosio^73^, Enrico Martinelli^73^, Sandro Mancarella^74^, Luisa Tavecchia^74^, Lia Crotti^75,76,77, 78^

22) Dept of Specialized and Internal Medicine, Tropical and Infectious Diseases Unit, Azienda Ospedaliera Universitaria Senese, Siena, Italy

23) Unit of Respiratory Diseases and Lung Transplantation, Department of Internal and Specialist Medicine, University of Siena, Italy

24) Dept of Emergency and Urgency, Medicine, Surgery and Neurosciences, Unit of Intensive Care Medicine, Siena University Hospital, Italy

25) Department of Medical, Surgical and Neuro Sciences and Radiological Sciences, Unit of Diagnostic Imaging, University of Siena, Italy

26) Rheumatology Unit, Department of Medicine, Surgery and Neurosciences, University of Siena, Policlinico Le Scotte, Italy

27) Department of Specialized and Internal Medicine, Infectious Diseases Unit, San Donato Hospital Arezzo, Italy

28) Dept of Emergency, Anesthesia Unit, San Donato Hospital, Arezzo, Italy

29) Department of Specialized and Internal Medicine, Pneumology Unit and UTIP, San Donato Hospital, Arezzo, Italy

30) Department of Emergency, Anesthesia Unit, Misericordia Hospital, Grosseto, Italy

31) Department of Specialized and Internal Medicine, Infectious Diseases Unit, Misericordia Hospital, Grosseto, Italy

32) Clinical Chemical Analysis Laboratory, Misericordia Hospital, Grosseto, Italy

33) Department of Preventive Medicine, Azienda USL Toscana Sud Est, Italy

34) Territorial Scientific Technician Department, Azienda USL Toscana Sud Est, Italy

35) Clinical Chemical Analysis Laboratory, San Donato Hospital, Arezzo, Italy

36) Chirurgia Vascolare, Ospedale Maggiore di Crema, Italy

37) Department of Health Sciences, Clinic of Infectious Diseases, ASST Santi Paolo e Carlo, University of Milan, Italy

38) Division of Infectious Diseases and Immunology, Fondazione IRCCS Policlinico San Matteo, Pavia, Italy

39) Department of Internal Medicine and Therapeutics, University of Pavia, Italy

40) Department of Anesthesia and Intensive Care, University of Modena and Reggio Emilia, Modena, Italy

41) Department of Medical and Surgical Sciences for Children and Adults, University of Modena and Reggio Emilia, Modena, Italy

42) HIV/AIDS Department, National Institute for Infectious Diseases, IRCCS, Lazzaro Spallanzani, Rome, Italy

43) III Infectious Diseases Unit, ASST-FBF-Sacco, Milan, Italy

44) Department of Biomedical and Clinical Sciences Luigi Sacco, University of Milan, Milan, Italy

45) Infectious Diseases Clinic, Department of Medicine 2, Azienda Ospedaliera di Perugia and University of Perugia, Santa Maria Hospital, Perugia, Italy

46) Infectious Diseases Clinic, “Santa Maria” Hospital, University of Perugia, Perugia, Italy

47) Department of Infectious Diseases, Treviso Hospital, Local Health Unit 2 Marca Trevigiana, Treviso, Italy

48) Clinical Infectious Diseases, Mestre Hospital, Venezia, Italy.

49) Infectious Diseases Clinic, ULSS1, Belluno, Italy

50) Department of Molecular Medicine, University of Padova, Italy

51) Department of Infectious and Tropical Diseases, University of Brescia and ASST Spedali Civili Hospital, Brescia, Italy

52) Department of Molecular and Translational Medicine, University of Brescia, Italy; Clinical Chemistry Laboratory, Cytogenetics and Molecular Genetics Section, Diagnostic Department, ASST Spedali Civili di Brescia, Italy

53) Medical Genetics and Laboratory of Medical Genetics Unit, A.O.R.N. “Antonio Cardarelli”, Naples, Italy

54) Department of Molecular Medicine and Medical Biotechnology, University of Naples Federico II, Naples, Italy

55) CEINGE Biotecnologie Avanzate, Naples, Italy

56) IRCCS SDN, Naples, Italy

57) Unit of Respiratory Physiopathology, AORN dei Colli, Monaldi Hospital, Naples, Italy

58) Division of Medical Genetics, Fondazione IRCCS Casa Sollievo della Sofferenza Hospital, San Giovanni Rotondo, Italy

59) Department of Medical Sciences, Fondazione IRCCS Casa Sollievo della Sofferenza Hospital, San Giovanni Rotondo, Italy

60) Clinical Trial Office, Fondazione IRCCS Casa Sollievo della Sofferenza Hospital, San Giovanni Rotondo, Italy

61) Department of Health Sciences, University of Genova, Genova, Italy

62) Infectious Diseases Clinic, Policlinico San Martino Hospital, IRCCS for Cancer Research Genova, Italy

63) Microbiology, Fondazione Policlinico Universitario Agostino Gemelli IRCCS, Catholic University of Medicine, Rome, Italy

64) Department of Laboratory Sciences and Infectious Diseases, Fondazione Policlinico Universitario A. Gemelli IRCCS, Rome, Italy

65) Department of Cardiovascular Diseases, University of Siena, Siena, Italy

66) Otolaryngology Unit, University of Siena, Italy

67) Department of Internal Medicine, ASST Valtellina e Alto Lario, Sondrio, Italy

68) Study Coordinator Oncologia Medica e Ufficio Flussi Sondrio, Italy

69) First Aid Department, Luigi Curto Hospital, Polla, Salerno, Italy

70) Local Health Unit-Pharmaceutical Department of Grosseto, Toscana Sud Est Local Health Unit, Grosseto, Italy

71) U.O.C. Laboratorio di Genetica Umana, IRCCS Istituto G. Gaslini, Genova, Italy.

72) Infectious Diseases Clinics, University of Modena and Reggio Emilia, Modena, Italy.

73) Department of Respiratory Diseases, Azienda Ospedaliera di Cremona, Cremona, Italy

74) U.O.C. Medicina, ASST Nord Milano, Ospedale Bassini, Cinisello Balsamo (MI), Italy

75) Istituto Auxologico Italiano, IRCCS, Department of Cardiovascular, Neural and Metabolic Sciences, San Luca Hospital, Milan, Italy.

76) Department of Medicine and Surgery, University of Milano-Bicocca, Milan, Italy

77) Istituto Auxologico Italiano, IRCCS, Center for Cardiac Arrhythmias of Genetic Origin, Milan, Italy.

78) Istituto Auxologico Italiano, IRCCS, Laboratory of Cardiovascular Genetics, Milan, Italy

### Spanish COVID Human Genetic Effort

Sergio Aguilera-Albesa^79^, Sergiu Albu^80^, Carlos Casasnovas^81,13^, Valentina Vélez-Santamaria^81,13^, Juan Pablo Horcajada^82^, Judit Villar^82^, Agustí Rodríguez-Palmero^83,13,14^, Montserrat Ruiz^13,14^, Luis M Seijo^84^, Jesús Troya^85^, Juan Valencia-Ramos^86^, Marta Gut^87^

79) Navarra Health Service Hospital, Pamplona, Spain

80) Institut Guttmann Foundation, Badalona, Barcelona, Spain

81) Bellvitge University Hospital, L’Hospitalet de Llobregat, Barcelona, Spain

82) Hospital del Mar, Parc de Salut Mar, Barcelona, Spain

83) University Hospital Germans Trias i Pujol, Badalona, Barcelona, Spain

84) Clínica Universitaria de Navarra, Madrid, Spain

85) Infanta Leonor University Hospital, Madrid, Spain

86) University Hospital of Burgos, Burgos, Spain

87) CNAG-CRG, Centre for Genomic Regulation (CRG), Barcelona Institute of Science and Technology (BIST), Carrer Baldiri i Reixac 4, 08028, Barcelona, Spain

